# The effect of music interventions in autism spectrum disorder: A systematic review and meta-analysis

**DOI:** 10.1101/2025.07.03.25330837

**Authors:** Laura Navarro, Nour El Zahraa Mallah, Wiktor Nowak, Jacobo Pardo-Seco, Alberto Gómez-Carballa, Sara Pischedda, Sensogenomics Working Group; Federico Martinón-Torres, Antonio Salas

**Affiliations:** Unidade de Xenética, Instituto de Ciencias Forenses, Facultade de Medicina, Universidade de Santiago de Compostela, and Genética de Poblaciones en Biomedicina (GenPoB) Research Group, Instituto de Investigación Sanitaria (IDIS), 15706 Hospital Clínico Universitario de Santiago (SERGAS), Galicia, Spain; Genetics, Vaccines and Infections Research Group (GenViP), Instituto de Investigación Sanitaria de Santiago, 15706 Universidade de Santiago de Compostela, Santiago de Compostela, Galicia, Spain; Centro de Investigación Biomédica en Red de Enfermedades Respiratorias (CIBER-ES), Madrid, Spain; Translational Pediatrics and Infectious Diseases, Department of Pediatrics, 15706 Hospital Clínico Universitario de Santiago de Compostela, Santiago de Compostela, Galicia, Spain

**Keywords:** autism spectrum disorder, neurodegeneration, neurodevelopment, music-based interventions

## Abstract

Several disciplines have approached the relationship between autism spectrum disorder (ASD) and music, but most of this understanding comes from cognitive sciences. This complex relationship has been studied by exploring how music-based interventions (MI) can benefit individuals with ASD. This systematic review and meta-analysis synthesize a range of evidence regarding the therapeutic effects of music on different aspects, including communication, behavior, social engagement, attention, and quality of life for those with ASD. Additionally, it contextualizes these effects within current research on the musical perception and processing abilities of ASD individuals, emphasizing how they perceive and process music. The studies reviewed employ a variety of methodologies, from randomized controlled trials to qualitative research, showcasing a wide array of interventions such as active music-making, music listening, and improvisational techniques. Despite substantial heterogeneity across studies, the findings point to a moderate overall benefit of MI, particularly in areas such as social interaction, expressive language, and quality of life. Given the evidence supporting the context-sensitive and domain-specific benefits of musical abilities in individuals with ASD, along with the positive outcomes highlighted in various studies, we conclude that music represents a valuable therapeutic tool for ASD. It engages individuals on emotional, cognitive, and social levels, providing a non-invasive and enjoyable way to enhance therapeutic outcomes. Future research should focus on individual differences, harmonization of outcome measures, and long-term effectiveness, paving the way for more personalized and neurodiversity-affirming intervention models.

## 1. Introduction

Autism Spectrum Disorder (ASD) is a complex neurodevelopmental condition characterized by altered perception [1; 2], sensory processing difficulties [3; 4], repetitive behaviors, rigid routines, a strong preference for sameness, an intense interest in specific topics or activities, and ongoing challenges in social communication [5]. ASD children often show a variety of unusual behaviors during their early years, compared to typically developing (TD) children, including resistance, repetitive actions, irritability, social withdrawal, reduced engagement, stereotyped behaviors, and atypical speech patterns. This might help explain why they struggle to integrate into society and build positive peer and family relationships, and why they often lack the social skills needed for everyday social situations.

Autism can profoundly affect both children and their parents, impacting daily life, finances, physical health, and mental well-being [6]. Due to the complex nature of ASD, which involves a mix of developmental and environmental factors and genetics, there is an ongoing debate about how the current treatments for ASD manage behavioral symptoms. Some interventions that have been used in the past include Applied Behavioral Analysis (ABA) [7; 8], Cognitive Behavioral Therapy (CBT) [9; 10], sensory integration training [11], and pharmacological treatments [12]. However, these approaches often require a long treatment period, and since every child with ASD is unique, the safety and effectiveness of these therapies are not yet fully supported by robust evidence [13]. Moreover, many of these interventions are limited in number and lack robust empirical validation. Therefore, it is crucial to identify scientifically sound and effective therapeutic approaches that can alleviate certain symptoms in children with ASD and improve their behavioral outcomes.

Music-based interventions (MI), such as singing, playing instruments, music listening, and music therapy, are one of the therapeutic approaches that have been gaining interest over the past few decades, as a way to support ASD individuals. Such interventions are notable for their connection with people through verbal and non-verbal modalities. These approaches tap into the unique qualities of music to achieve a range of developmental goals, such as enhancing communication [14; 15], encouraging emotional expression [16; 17], improving social interaction [15; 18], and boosting the overall quality of life (QoL) [18] for autistic individuals. Consequently, these accessible and cost-effective interventions highlight music’s universal appeal and its capacity to engage multiple domains of functioning, including cognitive, emotional, and social processes.

The benefits of music go beyond its fundamental neurological effects and play a significant role in neuro-rehabilitation practices [19]. Neuroscience research has uncovered several links between music and autism, particularly highlighting the distinct cognitive and sensory processing traits found in ASD individuals [20; 21; 22; 23]. For instance, neuroimaging studies have revealed that music activates different brain regions related to auditory processing in people with ASD compared to TD. A study [22] utilizing functional Magnetic Resonance Imaging (fMRI) to compare how the brains of individuals with and without autism respond to music has indicated that those with ASD exhibited greater activation in brain areas linked to music perception, such as the primary auditory cortex and regions associated with emotional processing. This suggests that music may have a more pronounced effect on brain networks in individuals with ASD.

Although numerous studies have examined the effects of MI on various outcomes in individuals with ASD, the results remain highly variable and at times contradictory. Variability in study design, participant characteristics, intervention types, and outcome measures has contributed to these inconsistencies. To address this, our review seeks to clarify the existing evidence regarding the musical abilities and perceptual processing of autistic individuals (Section 3.1), highlighting effective musical practices that contribute to a deeper understanding of music as a valuable tool in both everyday life and therapeutic contexts for this population. Furthermore, by way of a meta-analysis, our investigation evaluates the current evidence on the efficacy of MI for individuals with ASD, with a particular focus on areas such as overall improvement, social interaction, communication, quality of life, attention, and behavioral regulation (Section 3.2).

## 2. Methodology

### 2.1. Systematic review

Our investigation has a dual approach: (*i*) to discover the relation between music and autism by analyzing the evidence regarding the musical processing of music of ASD individuals, and (*ii*) to provide a better understanding of how music influences different aspects of ASD by integrating insights from neuropsychology.

A systematic review was conducted using PubMed, Cochrane Library, and WILEY Online Library databases, with the search extending between January 1^st^, 2020, and February 28^th^, 2024. To ensure comprehensive results relevant to ASD, we only selected the search term “autism” based on its prevalence in the literature. The term “music” was purposely kept broad to avoid restricting the scope of the study. We used the [tiab] field tag (title and abstract) to narrow the search to articles containing these specified terms, effectively filtering out irrelevant papers. Hence, the search query used was: (“autism”[tiab]) AND (“music”[tiab]).

The initial search yielded 346 publications, which were subsequently refined by eliminating duplicates, systematic reviews, and meta-analyses using the databases’ filtering tools. The selection was further narrowed to include only English-language articles focused on human subjects, published between 2000 and 2024, to ensure the inclusion of recent and relevant studies. This process resulted in a total of 285 publications. Of these, 65 were excluded for being reviews or systematic analyses. Following a detailed evaluation of the remaining 220 articles, additional exclusions were made for studies that either employed mixed therapeutic approaches involving music or fell outside the scope of the intended research focus. After this thorough analysis, 115 publications met all inclusion criteria and were selected for the systematic review.

Additionally, a total of five additional relevant articles were manually added to the list by scrutinizing the reference list of the selected articles. With a total of 120 publications, the findings were categorized into two main categories: (*i*) the synthesis of musical processing in individuals with ASD and (*ii*) the effects of music-based interventions for individuals with ASD. The present review was undertaken and is reported following the Preferred Reporting Items for Systematic reviews and Meta-Analyses (PRISMA) guidelines (https://www.bmj.com/content/372/bmj.n71); **Figure 1**.

**Figure 1.**
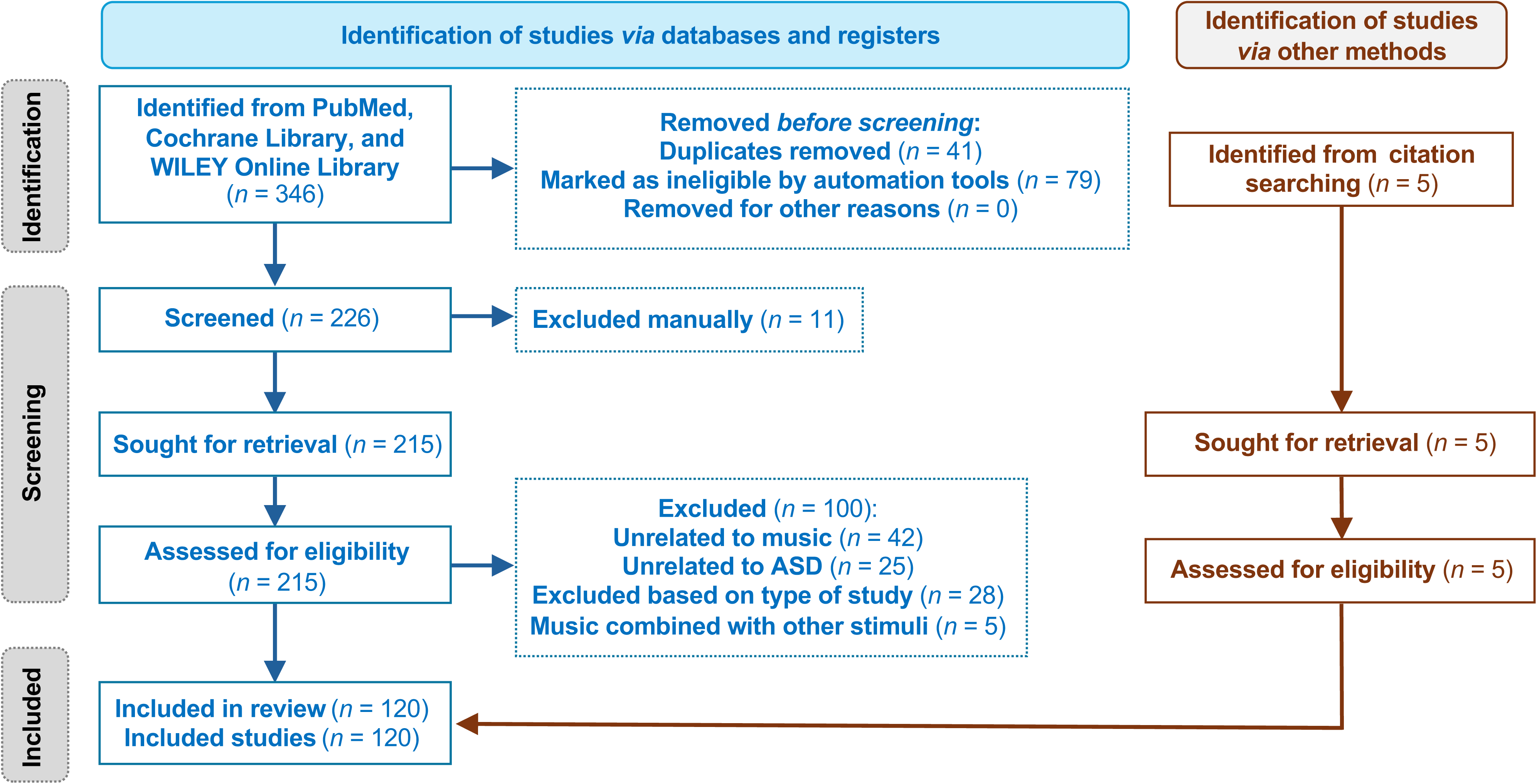
Flowchart of the study selection process following PRISMA guidelines. This visual representation illustrates how studies were identified, screened, assessed for eligibility, and ultimately included. The diagram details records retrieved via PubMed and citation tracking, as well as the number of studies excluded at each stage and the reasons for their exclusion. The structure follows the updated PRISMA model as outlined by Page et al. [119].

### 2.2. Meta-analysis

In addition to the systematic review and qualitative synthesis, this study includes a meta-analysis of 15 studies that met the criteria for quantitative evaluation, focusing on primary outcome domains commonly examined in ASD research. A structured and standardized methodology was employed to integrate findings across these outcome categories. Two reviewers independently extracted the relevant data, and discrepancies were resolved by consensus. Studies were excluded if they lacked a control group, exclusively reported short-term physiological markers, or failed to isolate the effect of MI from other co-occurring therapies. Data extracted from each study included sample sizes, means, and standard deviations for intervention and control groups, outcome measures used, intervention duration, and design features. Pooled mean differences and 95% confidence intervals (CIs) for pre-to-post changes were calculated for each domain, comparing MI groups with non-music controls. A random-effects model was applied to account for heterogeneity among studies, using the DerSimonian and Laird method [24]. Random-effects models are generally preferred when between-study heterogeneity is non-negligible, as was the case in several domains of this review, with I² values exceeding 50% [25]. Although this approach typically results in wider CIs due to the inclusion of between-study variance, it provides more conservative and generalizable estimates under conditions of heterogeneity. The diversity of intervention formats and participant profiles justified this modeling choice, which supports the broader applicability of the findings. Heterogeneity was assessed through the I^2^ statistic, *τ*^2^ (tau-squared), and Cochran’s Q test. In cases of substantial heterogeneity, subgroup analyses were conducted to identify potential sources, and statistical significance between subgroups was tested using chi-squared (*χ*^2^) comparisons. Forest plots were generated to visualize individual and combined effect sizes. Although the number of studies included was insufficient for formal publication bias testing (e.g., funnel plots or Egger’s test), sensitivity analyses were performed to evaluate the stability of results by excluding studies with extreme values or high risk of bias. Subgroup analyses were also stratified by specific measurement instruments (e.g., ADOS-SA, SRS, ASSP), recognizing the varied constructs captured by each outcome assessment. To estimate the potential variability of treatment effects in future research settings, prediction intervals were computed, considering both intra-and inter-study variance. One recurring challenge in ASD meta-analyses is the lack of reported standard deviations for change-from-baseline outcomes [26]. In accordance with Cochrane recommendations [27], missing SDs were imputed using a conservative correlation coefficient of *r* = 0.8, consistent with assumptions made in prior meta-analytic work [26; 28]. To standardize treatment effects, outcome directions were harmonized by inverting scores where necessary. In cases where different tools assessed the same domain, scores were transformed to a common scale to support comparability. Nevertheless, variability in scoring systems and measurement units meant that not all studies could be integrated into the same subgroup analysis, limiting some within-domain comparisons. Despite this, the overall consistency of results supports the robustness of the findings.

Separate meta-analyses were conducted for each major domain (e.g., communication, social engagement), ensuring interpretability and consistency of effect sizes.

All statistical analyses were conducted using the R meta package [25], with forest plots illustrating the results and the statistical significance of pooled effects assessed using z-tests.

## 3. Results

Based on the included studies’ results and our investigation’s goal, the studies were divided into two main categories: (*i*) synthesis of current research on musical processing in individuals with ASD, focusing on their perception and musical abilities and (*ii*) discovery of benefits and therapeutic effects of music for ASD individuals.

### 3.1. Musical processing and musical abilities in people with ASD. A musical phenotype in autism?

We analyzed 45 articles on how music’s structural aspects are processed in autistic populations (overall musically untrained), compared to TD children. We aimed at investigating whether specificities exist in the musical brains of people with ASD, especially in musical perception and emotion processing. The main analyzed categories were pitch processing (pitch discrimination and absolute pitch), auditory pleasantness, emotional processing, other musical domains, and the function of music in their lives.

#### 3.1.1. Pitch discrimination abilities

The ability to identify a specific pitch involves cognitive processes such as auditory perception and memory. In this section, we examined the nature of auditory mechanisms underlying musical recognition and assess whether individuals with ASD exhibit distinct abilities or processing patterns. This field is the main corpus of papers corresponding to musical abilities in ASD.

The main finding is the special sensitivity of individuals with ASD to pitch changes and their enhanced ability to discriminate musical parameters, providing evidence for preserved, or even superior, pitch processing in this population [29; 30; 31; 32; 33; 34; 35; 36; 37; 38]. According to Chowdhury et al. [35], auditory perception is related to non-verbal reasoning rather than verbal abilities in ASD and TD. Some studies connect ASD‘s ability to recognize different pitches to superior memory, including long-term melodic memory [30; 33] or working memory, and focused attention [39]. However, Heaton et al. [36] affirmed that ASD individuals perform better than TD controls on pitch identification despite being impaired in short-term memory. Some studies detect similar recognition of melodic contour in ASD and TD [40], and other research affirmed that auditory imagery is lower in ASD than in TD controls [41].

Many studies compared the effect of listening to music with listening to speech. Järvinen-Pasley and Heaton [42], and Heaton et al. [43] demonstrated that when speech is included in perception activity, the levels of accuracy in pitch discrimination of ASD individuals decrease. Also, DePape et al. [34] demonstrated that ASD individuals are more impaired in speech tasks than in music tasks. In this line, Lai et al. [23] compared the neural systems and showed that ASD individuals activated the left inferior frontal gyrus more than controls in song stimulation; however, the opposite happened with speech stimulation. Sharda et al. [44] found that functional frontotemporal connectivity was preserved during sung-word listening in ASD in contrast to speech-word, enhancing the importance of MI with this population. Other arguments present evidence of how ASD mental representations of pitch contours could be across domains, and implications for using music to improve language [45].

#### 3.1.2. Absolute pitch

Absolute pitch (AP) is an extreme phenotype associated with naming or producing a musical tone without any reference. Several researchers have suggested that AP is a normally distributed complex trait with a strong genetic component [46; 47], with a prevalence of <1% in the general population [48].

The outstanding study of DePape et al. [34] measured the prevalence of AP processing in ASD using a task that does not require explicit knowledge of musical structure, which can be used by non-musicians with and without ASD, estimating a prevalence of 11% in autism, which is remarkable compared to 1 in 10,000 in typical populations.

Autism‘s neurocognitive theories might explain this co-occurrence, while some case studies illustrate evidence of AP in individuals with ASD who possess extraordinary musical abilities [49; 50; 51; 52]. Research involving musicians has found autistic features in those with AP, suggesting a link between the two conditions [53; 54; 55; 56; 57] and highlighting the detail-oriented cognitive style, imagination, perceptual functioning, and hyper-systemizing present in both. A noteworthy study emphasizes a potential connection between autistic traits, brain connectivity, and AP ability [57], suggesting a less efficient and less small-world-structured functional network in AP, consistent with findings from autism research.

Curiously, one study focusing on pitch production reveals a vocal imitation deficit in individuals with ASD that is specific to AP, but not to relative pitch, across both speech and music domains [58]. This may be related to the fact that individuals with ASD often exhibit atypical imitation of actions and gestures.

#### 3.1.3. Other musical abilities

Pitch recognition is related to the ability to produce a determined sound properly. The processing involving human listening, specifically musical discrimination, is inevitably joined to musical production through intonation [59]. Regarding the auditory processing that involves pitch production through voice, Wang et al. [37] found that imitating musical intonation is intact in ASD individuals. Other studies compared this ability of pitch production in musical language with speech production, demonstrating the preservation of pitch production and a prosodic impairment [60; 61].

According to Heaton [30], abilities such as pitch memory and labeling are superior in ASD and may facilitate performance in harmonic contexts for ASD individuals. In a subsequent study, Heaton et al. [43] affirmed that the most striking finding was the absence of significant differences in performance patterns between ASD and control participants; both groups were similarly influenced by harmonic context in their perception. DePape et al. [34] found that musical processing is relatively preserved in ASD in many aspects: pitch detection, pitch memory, harmonic and metrical processing. Other minor examinations focused on rhythmic perception and affirmed that the tempo of acuity was preserved [36; 62].

#### 3.1.4. Auditory pleasantness

Auditory perception and overall musical pleasantness depend on both the acoustic properties of the stimulus and the cultural experiences that shape individual musical preferences. Six studies have investigated musical preferences in individuals with ASD, while others have focused on the benefits of different types of music and the importance of selecting appropriate musical content for therapeutic purposes. While recent research [63] shows ASD adults rated instrumental sounds more pleasant than vocal sounds, Kalas [64] discovered the effectiveness of joint attention of simple music (short melody without syncopation or chromatism, accompanied by I-IV-V chords) for severe ASD, in contrast to the function of complex music for mild/moderate ASD people. In qualitative research [65], parents of ASD children reported an active response to music and a preference for rhythmic music. All research compared the auditory pleasantness of ASD people with controls, reaching relevant results as ASD are more sensitive to consonance and dissonance, appreciating a larger variety of music, from Mozart to Schoenberg [66]. The most compelling study examines functional brain connectivity between specific regions while comparing familiar and unfamiliar songs [67], concluding that there is no difference in how ASD and TD individuals process familiar music.

However, this author found significant differences in how the ASD brain processes unfamiliar music compared to the TD, activating the alpha band and increasing connectivity. Also, in this research, ASD children showed no difference in the magnetoencephalography of familiar and unfamiliar music. However, Lanovaz et al. [68] found that preferred music reduced vocal stereotypy in four ASD children.

#### 3.1.5. Emotional processing of music

Lately, processing musical emotions in individuals with ASD, such as emotion recognition or emotional expressions, has gained attention. The impairment to identify, recognize, or verbalize emotions is known as alexithymia, but ASD individuals could be affected only in the ability to verbalize or articulate the expression of emotions [69; 70]. Music improves emotion recognition in ASD individuals [21; 71], but many studies contribute deeply to understanding the mechanisms underlying emotional processing.

Some studies demonstrated the physiological effect of music on the ASD population by measuring the emotional response through skin conductance or the autonomic nervous system‘s reaction, or evaluating the capability to recognize emotions, demonstrating no notable difference between ASD and controls [17; 70; 72; 73; 74], and the accuracy in emotional recognition in ASD individuals [72; 74; 75].

Also, using fMRI, neuroscientific studies found similar neural networks during musical processing in emotion recognition in ASD and TD [76; 77], considering music as a domain of preserved ability. However, Caria et al. [76] associated the strong emotional response to happy music in ASD with decreased activity in the cerebellum and premotor areas, showing a possibly altered rhythm perception.

However, some studies found an impairment in judging the emotional expressivity of music [78] or established differences between ASD and TD in the emotional response to music. Quintin et al. [17] studied the recognition of happiness, sadness, scariness, and peacefulness through 20 musical clips, and they only found a difference between ASD and TD adolescents in recognition of the peaceful music, which was the most difficult. Gebauer (2014) (PMID: 25076869) showed that ASD individuals demonstrate more arousal activity and cognitive load in happy than in sad music, unlike TD controls.

Leung et al. [74] found that the processing speed of emotions through music was slower in the ASD group compared to the NT group, suggesting that individuals with ASD may employ different emotion-processing strategies. Also, Wagener et al. [21] found higher reaction times in ASD. Stephenson et al. [79] found reduced skin conductance in response to music in ASD adolescents, as well as a decrease in physiological responsiveness with age, contrary to NT controls.

#### 3.1.6. Qualitative Role of Music

Several studies employing qualitative or mixed-method designs have explored the meanings and functions of music in ASD peoplés lives. For example, Kirby and Burland [80] identified four primary roles that music plays: emotional, cognitive, identity-related, and social. Notably, they found these roles to be consistent across ASD and TD individuals. Importantly, autistic adults often use music for emotional self-regulation; they engage with music to modulate mood, manage emotional states, and support affect regulation [69; 81; 82]. Furthermore, music also serves as a means of fostering social connection and interaction [81; 82], highlighting its potential as a bridge for communication and social engagement in ASD populations.

### 3.2. Music-Based Interventionś Impact on ASD

Music-based interventions (MI) can play a crucial role in improving people’s lives, demonstrating the potential of music as a valuable therapeutic tool for supporting ASD individuals. In the 75 studies included in this systematic review, 34 utilized a Music Therapy (MT) approach and 41 a different music-based intervention, of which 17 are based on listening to music, 7 on playing musical instruments, 6 on singing and create songs, 5 on music and movement intervention, and 6 in combining musical activities. All of the musical interventions reported benefits, but especially an active engagement, such us MT, playing instruments, music and movement or a combination of playing, singing and improvisation can effectively minimize core symptoms in autistic children and can help in reducing autism severity, enhancing social engagement, and decreasing repetitive behaviors, among others [83].

Many different techniques of MI have been utilized in the 34 studies reviewed, such as Receptive Music Therapy, Improvisational Music Therapy, or Neurological Music Therapy, among others, impacting especially in social engagement and behavioral outcomes (see for example [84]). In a comparative study, Rabeyron et al. [85] found that MT led to greater clinical improvements, measured using the Clinical Global Impression (CGI) scale, and greater reductions in stereotypical behaviors than music listening (ML) alone. Notably, MT was associated with enhanced social engagement in 27 studies, whereas other music-based interventions reported similar benefits in 22 studies. Behavioral improvements were observed across various musical activities, with 11 MT studies and 9 ML interventions specifically highlighting such effects.

Next, we provide a detailed summary of the findings of the 15 studies that fulfilled the inclusion criteria for meta-analysis, out of the 120 articles included, focusing on verbal and non-verbal communication, attention, behavior, QoL, and social interaction. **Table 1** details the outcomes and the measures of these included studies.

**Table 1.** Overview of studies included in the systematic review, including intervention duration and outcome assessment tools. Age expressed in years. Abbreviations: A: Attention; B: Behavior; GI: Global Improvement; nVC: Non-verbal Communications; QoL: Quality of Life; SI: Social Interaction; VC: Verbal Communication.

| Reference | Age | Outcomes | Measures |
| --- | --- | --- | --- |
| Bieleninik et al. (2017) | 4-7 | GI/QoL/SI | Autism Diagnostic Observation Schedule-Social Affect (ADOS-SA)<br>Social Responsiveness Scale (SRS) Total<br>ADOS-Restricted and Repetitive Behaviors (ADOS-RRB)<br>Parent-reported QoL of the child (QoL-child) and of the family as a whole (QoL-family) |
| Chenausky et al. (2022) | 5-11 | VC | % Vowels Correct (%Vc)<br>% Syllables Approximately Correct (%Sac)<br>% Consonants Correct (%Cc) |
| Gattino et al. (2011) | 7-12 | G/nVC | Childhood Autism Rating Scale, Brazilian version (CARS-BR)<br>CARS-BR non-verbal communication domain<br>CARS-BR verbal communication domain |
| Ghasemtabar et al. (2015) | 7-12 | SI | Social Skills Rating System Scale (SSRS) |
| Kim et al. (2008) | 3-5 | GI/SI | Social Approach Subscale (Pervasive Developmental Disorder Behavior Inventory, PDDBI)<br>Early Social Communication Scale (ESCS) |
| LaGasse et al. (2014) | 6-9 | GI/nVC/A | Social Responsiveness Scale (SRS) Total<br>Autism Treatment Evaluation Checklist (ATEC) Total |
| LaGasse et al. (2019) | 5-12 | B | Behavior measures |
| Lim et al. (2010) | 3-5 | VC | Verbal Production Evaluation Scale (VPES) |
| Ren et al. (2022) | 5-8 | B/GI | Childhood Autism Rating Scale (CARS)<br>Autism Treatment Evaluation Checklist (ATEC) |
| Sa et al. (2020) | 10-14 | A | Test of Everyday Attention for Children 2 (TEA-Ch 2) |
| Schwartzberg et al. (2013) | 9-21 | GI/SI/VC | Autism Social Skills Profile (ASSP)<br>Comprehension checks (CCs)" |
| Sharda et al. (2018) | 6-12 | B/nVC/QoL/SI /VC | Social Responsiveness Scale (SRS-2)<br>Beach Center Family Quality of Life Scale (FQoL)<br>Children's Communication Checklist (CCC-2)<br>Peabody Picture Vocabulary Test-4 (PPVT-4)<br>Vineland Adaptive Behaviour Scales (VABS) |
| Srinivasan et al. (2015) | 5-12 | B | Repetitive Behavior Scale (RBS) |
| Thompson et al. (2014) | 3-6 | GI/SI/VC | Parent-Child Relationship Inventory (PCRI)<br>Vineland Social Emotional Early Childhood Scales (Vineland SEEC)<br>Social Responsiveness Scale-Preschool version (SRS-PS)<br>MacArthur-Bates Communicative Development Inventories-Words and Gestures (MBCDI-W&G) |
| Williams et al. (2024) | 2-5 | B/SI/VC | Social Responsiveness Scale (SRS-2)<br>Expressive One-Word Picture Vocabulary Test (EOWPVT-4)<br>Receptive One-Word Picture Vocabulary Test (ROWPVT-4)<br>MacArthur-Bates Communicative Development Inventories (CDI)<br>Vineland Adaptive Behaviour Scales-third edition (VABS-3)" |

#### 3.2.1. Attention

ASD is characterized by differences in attention regulation, which can influence cognitive processing and sensory perception. Music has emerged as a potential tool to manage attention in ASD people. It engages both auditory and emotional networks, which can lead to better focus and less distractibility. A study [86] examined how autistic and non-autistic young adult drivers performed while listening to music in various situations, and demonstrated that music might have distinct effects on attention and task performance in ASD individuals compared to TD. Additionally, a randomized controlled trial [87] investigated how parent-child collaborative MT could benefit ASD children and their mothers, indicating potential improvements in attention for children as well as improved maternal well-being. Moreover, Kim et al. [88] reported that ASD children who engaged with music demonstrated better attention than those who did not. Also, Pasiali et al. [89] highlighted the potential of MI in advancing attention in autistic individuals, emphasizing how engaging and motivating music can be. Collectively, these studies underscore the significant role of music in enhancing attention in autism, suggesting that personalized MI might be helpful in both therapeutic and everyday contexts.

Structured MI, like rhythmic entrainment and melodic stimuli, might enhance attentional control and improve task performance ASD population as well. LaGasse et al. [84] provided evidence that rhythmic training through music can improve joint attention between autistic children. Furthermore, Sa et al. [90] supported these findings by showing that structured musical activities can facilitate attentional shifts and reduce distractibility in those with ASD.

We conducted a meta-analysis to evaluate the impact of MI on attention in individuals with ASD, using the Test of Everyday Attention for Children (TEA-Ch), the Red & Blues, Bags & Shoes (RBBS), and Joint Attention tests as primary outcome measures (**Figure 2**). The analysis included two methodologically sound studies [90; 91] and employed a random-effects model to pool their results. The overall mean difference was 1.2 with a 95% confidence interval (CI) of [−6.09; 8.49], but the test for overall effect was not statistically significant (z = 0.32, *P*-value = 0.75). Heterogeneity was substantial (I^2^ = 77%, *τ*^2^ = 55.30, *P*-value < 0.01), indicating considerable variation in findings across studies. The studies in this meta-analysis contributed to comparable statistical weights, ranging from 17.6% to 24.6%. It is important to note the outlier behavior observed in one of the measurements reported by Sá et al. [90]; **Figure 2**. When this outlier is excluded from the meta-analysis, the overall metrics improve notably (**Figure S1**). Specifically, the pooled mean difference increases to 3.66 (95% CI: −2.49 to 9.80). Although the overall effect remains statistically non-significant, the test statistic shows a stronger trend (z = 1.17, *P* = 0.24). In addition, heterogeneity decreases substantially (I^2^ = 67%, *τ*^2^ = 26.60, *P*-value = 0.03), though considerable variability across studies still persists.

**Figure 2.**
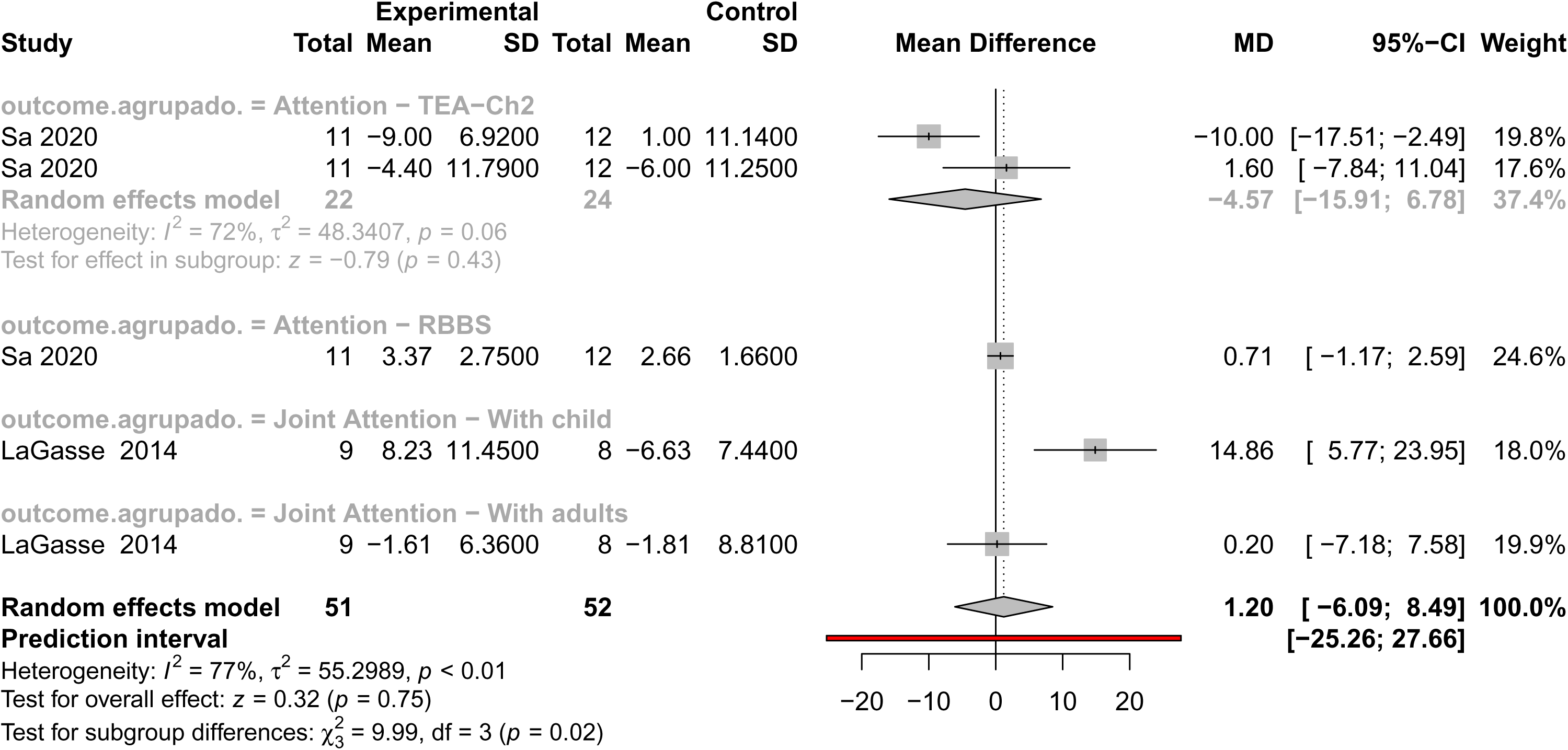
Forest plot illustrating the effect sizes (mean differences) for studies comparing interventions and controls about attention outcomes in individuals with ASD. The plot includes each study’s mean difference (MD), 95% confidence interval (95% CI), and statistical weight. Summary estimates were calculated using a random effects model. Heterogeneity statistics (I², τ², and p-values), prediction intervals, and tests for subgroup and overall effects are also provided.

Although the overall effect was not statistically significant, the mean difference of 1.2 suggests a positive trend favoring MI. However, the wide confidence interval and lack of statistical significance indicate that this trend should be interpreted with caution. Therefore, given the limited number of studies and small sample sizes, further research is necessary to clarify the potential of music to improve attentional functioning in this population, particularly concerning specific subtypes of attention.

#### 3.2.2. Behavior

ASD is marked by unique differences in how individuals process sensory information and behave, often showing repetitive patterns and difficulty with managing emotions. Music has been widely explored as an intervention to support behavioral and emotional growth in ASD individuals. Research suggests that MI can boost social engagement, ease anxiety, and improve adaptive behaviors by taking advantage of the structured, predictable, and emotionally expressive nature of music.

Recent studies have investigated how these MIs affect behavior in autistic individuals. Williams et al. [92] examined the use of MT by autistic adolescents and reported improvements in social interactions and a drop in anxiety levels. Moreover, Tahmazian et al. [93] investigated how rhythmic auditory stimulation impacts repetitive behaviors in ASD children and showed a notable decrease in these behaviors following the intervention. Another piece of research from Liu et al. [94] examined how musical activities can help adults with ASD manage their emotions. It turned out that regularly participating in music-based activities improved emotional control and reduced aggressive behaviors.

Expanding upon the review, Lundqvist et al. [95] investigated the use of a robot-based music therapy (MT) platform to enhance social behaviors in autistic children. Their findings indicated that most participants with ASD exhibited stable turn-taking behavior during musical activities. Notably, vibroacoustic music was found to reduce challenging behaviors and lower the incidence of self-injurious actions. These results suggest that the robot-assisted MT platform could be a promising tool for enhancing fine motor skills and turn-taking abilities in children with ASD. Overall, these insights support the notion that MI may positively influence various behavioral aspects in individuals with ASD, including repetitive behaviors, social skills, anxiety, and emotional regulation.

In the meta-analysis of behavioral outcomes, only five studies were included, with pooling data across studies using a random-effects model (**Figure 3**). The analysis incorporated several behavioral assessment tools, such as the Childhood Autism Rating Scale (CARS), Vineland Adaptive Behavior Scales (VABS), and Repetitive Behavior Scale (RBS). The overall test for effect yielded a mean difference that was not statistically significant (z = 1.61, *P*-value = 0.11), suggesting that the impact of music on general behavior, as currently measured, remains inconclusive. Importantly, heterogeneity across studies was extremely high (I^2^ = 98%, *τ*^2^ = 90.41, *P*-value < 0.01), indicating considerable variability in outcomes, populations, intervention types, and measurement instruments. Subgroup analyses revealed significant differences across behavioral scales (*χ*^2^ = 289.04, df = 7, *P*-value < 0.01), emphasizing that the magnitude and direction of effects may depend heavily on the specific behavioral domain assessed and the tool used. While some individual studies showed large positive effects (e.g., [92]), others reported mixed or null results, reflecting the complexity of measuring behavioral change in ASD through MI.

**Figure 3.**
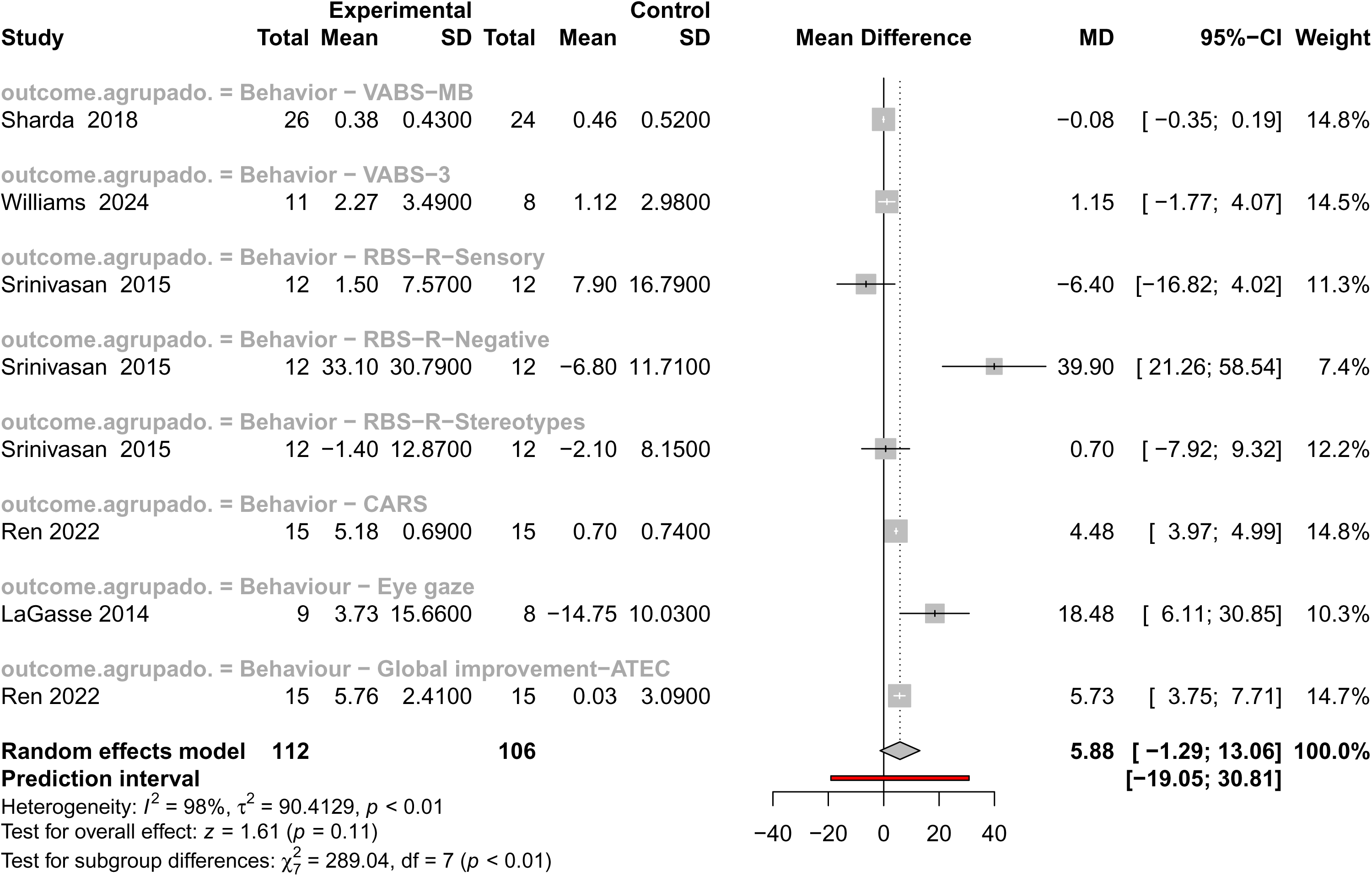
Forest plot depicting comparative mean differences for behavioral outcomes between intervention and control groups in individuals with ASD. The plot presents mean differences, 95% confidence intervals, and weights for each study. Summary estimates were calculated using a random effects model, along with heterogeneity measures (I², τ², and p-values), prediction intervals, and subgroup/overall test results.

Eliminating the outlier outcome represented by the RBS-R-Sensory measure from Srinivasan et al. [96] significantly improves the overall metrics (**Figure S2**). The effect estimate becomes more statistically robust, reaching marginal significance (z = 1.92, *P*-value = 0.06), suggesting a potential positive impact of music on general behavior. However, heterogeneity across studies remains extremely high (I^2^ = 98%, *τ*^2^ = 89.74, *P*-value < 0.01), indicating substantial variability in study results.

This meta-analysis provides suggestive statistical evidence of behavioral improvements. The observed trends and scale-specific heterogeneity indicate that MI may benefit certain behavioral aspects in individuals with ASD. These findings warrant further investigation using standardized protocols, clearly defined behavioral outcomes, and larger sample sizes.

#### 3.2.3. Communication

Communication challenges have always been considered a fundamental feature of autism, yet there is a vast range of differences in how people with autism communicate and express themselves. This variation not only highlights the unique nature of the condition but also the complexity of communication itself. It involves not just the chosen words and their sequence, but also eye contact, facial expressions, gestures, and other nonverbal signals. Because of this complexity, music has gained attention as a potential means to enhance communication for those with ASD. It taps into shared neural pathways that are crucial for emotional processing and social engagement. Studies suggest that MI, such as improvisational music therapy [20; 88; 97] and rhythmic entrainment [84; 96; 98], can enhance expressive abilities in autistic individuals. Moreover, the structured and predictable nature of music can create a comforting environment for communication, reduce anxiety, and encourage participation.

Another important connection between autism and music lies in language and verbal communication. Many ASD individuals face language challenges, such as delayed speech and unusual language development [5]. Music, especially its rhythmic and melodic elements, has been found to aid in language and development [15; 99]. The rhythmic patterns found in music reflect the natural rhythms of speech, which can improve vocabulary and sentence structure in ASD children, making the learning process more effective and enjoyable.

Based on the previously mentioned findings, both verbal and non-verbal communications were examined in our investigation.

##### 3.2.3.1. Verbal communication

ASD affects cognitive processing and communication, often impacting learning styles and sensory experiences. Music is widely acknowledged as a powerful tool for autistic individuals, offering a structured yet adaptable medium that supports cognitive, emotional, and social development. Research suggests that music enhances learning by improving attention, memory, and language skills while reducing anxiety and sensory overload [88; 100]. The rhythmic and repetitive structure of music appears to align particularly well with the cognitive and sensory profiles of individuals on the autism spectrum, enhancing both engagement and comprehension [101; 102]. Furthermore, MI, such as MT and adaptive music education, has shown success in fostering communication and self-expression.

The research by Schwartzberg et al. [103] may inspire music therapists and educators to integrate music-based stories into reading programs to improve comprehension skills in ASD children. Although this study did not reveal significant differences between the singing and reading groups, it underscored the potential of MI to aid reading comprehension in ASD children. Furthermore, the findings emphasized the necessity of employing diverse instructional methods for teaching reading comprehension, suggesting that music can be a valuable element of a comprehensive educational strategy for ASD children.

Various studies highlighted the effectiveness of music-based approaches in both therapeutic and diagnostic contexts for ASD individuals [104; 105] and have noted that MI not only improves motor skills in autistic children but also serves as a diagnostic tool, especially for populations with intellectual disabilities. Therefore, integrating music into learning environments can yield fruitful benefits for autistic people, fostering their development and overall well-being.

To evaluate the impact of MI on verbal communication in autism, we performed a meta-analysis by compiling data from six studies using various outcome measures, including Peabody Picture Vocabulary Test–Fourth Edition (PPVT-4), the Expressive One-Word Picture Vocabulary Test–Fourth Edition (EOWPVT-4), and Conversational Communicative Scales (CCs); **Figure 4**. The overall pooled analysis using a random-effects model yielded a non-significant effect (z = 0.66, *P*-value = 0.51), suggesting no consistent benefit of MI on verbal communication across the entire dataset. Importantly, there was substantial heterogeneity among the studies (I^2^ = 62%, *τ*^2^ = 0.04, *P*-value < 0.01), suggesting that true differences between study outcomes, beyond random sampling error, exist to a notable degree. A test for subgroup differences (*χ*^2^ = 15.83, df = 14, *P*-value = 0.32) was not statistically significant, but the result still indicated that the impact of MI may vary depending on the specific verbal communication outcome measured. While most subgroups showed no statistically significant changes, some specific tools, such as the Verbal Production Evaluation Scale (VPES), which focuses on the quantity and quality of verbal output, showed a moderately positive effect, though this result also did not reach statistical significance (z = 1.39, *P*-value =0.16). Even more notably, the “Vowels” subgroup demonstrated a stronger positive effect, with a higher standardized mean difference, reaching statistical significance (z = 2.93, *P*-value < 0.01; I^2^ = 0%, *τ*^2^ = 0, *P*-value = 0.83). This suggests that MI may be particularly effective in enhancing phonemic-level expressive language abilities, such as vowel articulation, in individuals with ASD.

**Figure 4.**
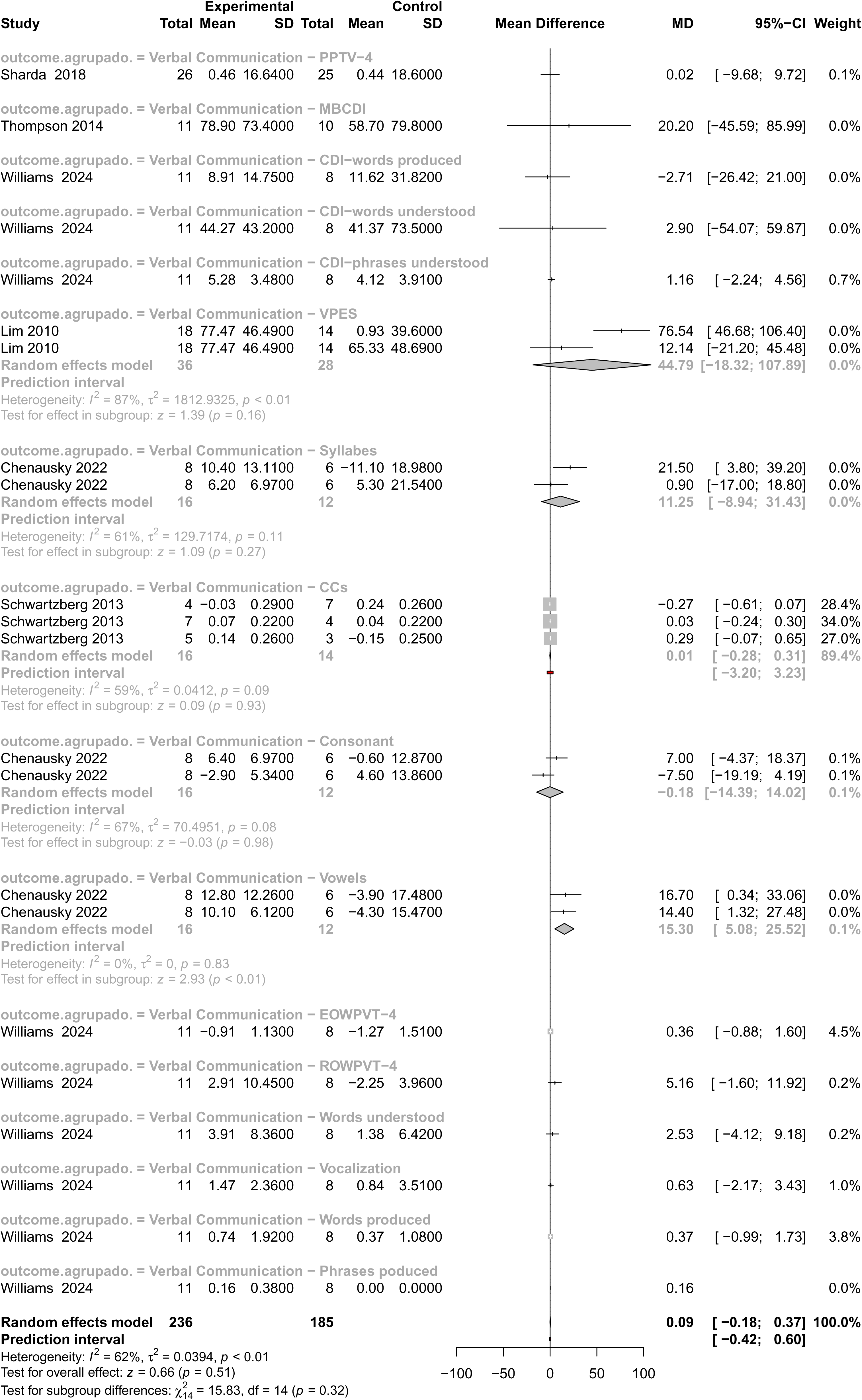
Forest plot summarizing intervention effects on verbal communication in individuals with ASD. Individual study data include mean differences, 95% confidence intervals, and weights, with pooled estimates computed using a random effects model. Heterogeneity measures, prediction intervals, and subgroup analyses are also presented.

This finding suggests that MIs may effectively enhance expressive language skills in individuals with ASD, as measured by the VPES. Notably, the heterogeneity within this subgroup was moderate to high (I^2^ = 87%, *τ*^2^ = 1812.93, *P*-value < 0.01), indicating a relatively consistent effect across the included studies, despite variability. A further breakdown across the entire meta-analysis revealed that heterogeneity within individual subgroups ranged from negligible (I^2^ = 0%, *τ*^2^ = 0) to moderate (e.g., I^2^ = 67%, *τ*^2^ = 70.50), highlighting that even within outcome-specific analyses, variability remains. These variations may be due to differences in intervention design (e.g., active music-making *vs*. passive listening), duration, therapist involvement, or participant characteristics such as age, language level, and ASD severity. Overall, while MI do not appear to produce a universal or robust effect on verbal communication in ASD when averaged across all studies, certain subsets of interventions or outcome measures may hold promise. The considerable heterogeneity highlights the complexity of music’s role in therapeutic settings for ASD, suggesting the need for more refined, targeted research designs and better harmonization of outcome measures in future studies.

Overall, although MI may not yield a universal or robust effect on verbal communication in ASD when results are averaged across all studies, promising trends in specific domains, such as expressive speech and vowel articulation, suggest meaningful potential for targeted therapeutic impact. The observed heterogeneity highlights not only the complexity of verbal outcomes in autism but also the opportunity to tailor interventions to individual profiles and specific communicative challenges. These findings reinforce the value of continuing research with more refined designs, outcome-specific analyses, and harmonized assessment tools to fully explore the capacity of music to support verbal development in individuals with ASD.

##### 3.2.3.2. Non-verbal communication

Communication impairments are considered a key diagnostic characteristic of ASD. Autistic people struggle with typical non-verbal communication methods such as facial expressions, gestures, body language, and maintaining eye contact, which can limit their overall communication skills [106; 107]. Music stands out as a fascinating, unique, and multi-faceted stimulus that engages our brain in processing different types of information simultaneously: visual, auditory, somatosensory, and motor. When individuals engage in music-making, they utilize these integrated sensory inputs to guide movement and expression [108]. Since music-making activities might engage regions of the brain that overlap with regions that presumably contain mirror neurons (engaged by seeing, hearing, and doing an action), recent research suggests that music, particularly structured interventions such as rhythm-based exercises [84; 96; 98], interactive singing [109], instrumental play [110], could provide a fun and effective way to boost non-verbal communication skills in autistic individuals. For instance, Sharda et al. [22] showed that an 8-to 12-week improvisational intervention not only enhanced social communication but also improved the connections between auditory and motor brain regions in ASD children.

Only three studies were eligible for inclusion in the meta-analysis evaluating the effect of MI on individuals with ASD **(Figure 5)**. The overall pooled effect size was small and statistically non-significant (MD = 1.82, 95% CI: −0.98 to 4.61, z = 1.28, *P*-value = 0.20), suggesting that MI did not lead to reliable improvements compared to control conditions. Heterogeneity was low (I^2^ = 38%, *τ*^2^ = 3.12, *P*-value = 0.20), indicating consistency across studies. The wide prediction interval (−27.02 to 30.66) suggests considerable uncertainty around the true effect under similar conditions. Among the included studies, Gattino et al. [111] contributed the most weight (64.5%), and used the Childhood Autism Rating Scale (CARS), reporting a small but statistically significant mean difference (MD = 0.60, 95% CI: 0.28 to 0.92). The study by LaGasse et al. [84] reported a more favorable mean difference (MD = 2.80), but a wide confidence interval (95% CI: −3.77 to 9.37). Similarly, Sharda et al. [22] also reported a slight improvement using the CCC-2 (MD = 4.84, 95% CI: −0.11 to 9.79), though with notable uncertainty.

**Figure 5.**
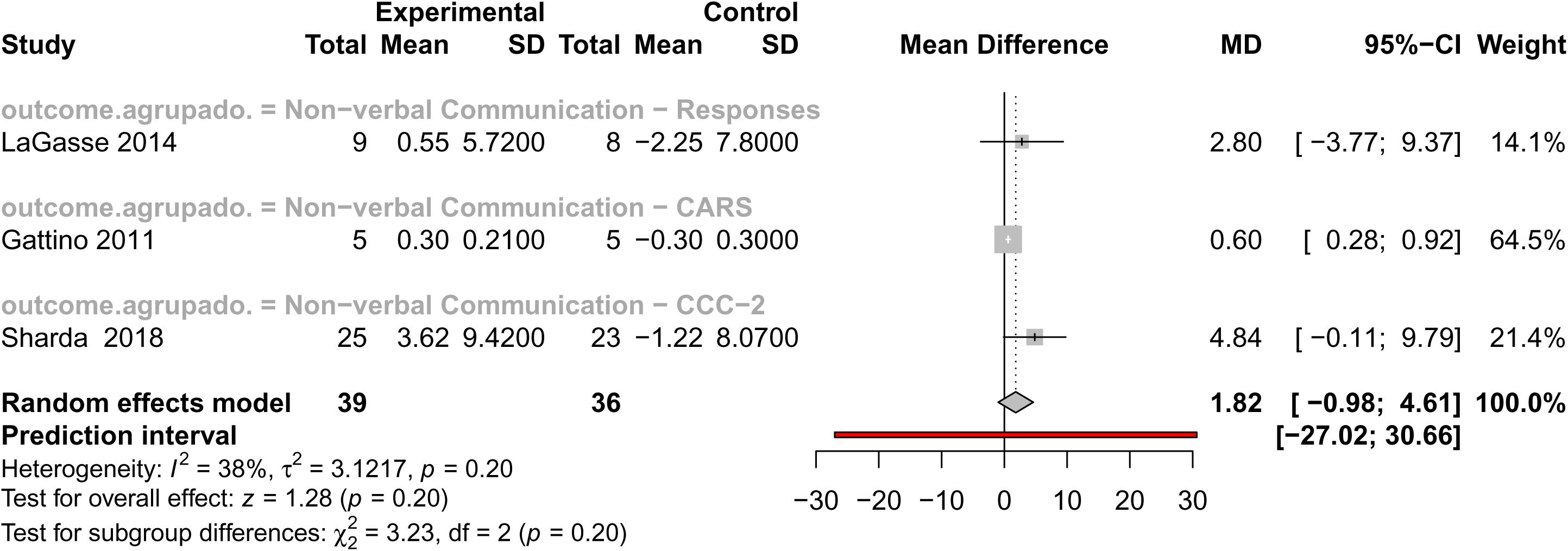
Forest plot displaying mean differences for non-verbal communication outcomes across studies involving individuals with ASD. Results include individual study MDs, confidence intervals, and weights, with overall estimates calculated via a random effects model. The plot also reports heterogeneity statistics, prediction intervals, and significance tests for subgroups and overall effects.

Taken together, while the current meta-analytic evidence does not confirm a consistent or statistically significant effect of MI on non-verbal communication in ASD, the presence of small positive trends, especially in studies using more sensitive or targeted measures, suggests potential. These findings highlight the need for further high-quality, large-scale studies to explore specific therapeutic conditions under which music may meaningfully benefit communication outcomes in individuals with ASD.

#### 3.2.4. Quality of life

Quality of life (QoL) is an essential factor in understanding ASD, as it includes physical, emotional, social, and psychological well-being. Autistic individuals often face challenges in communication, social engagements, sensory sensitivities, and mental health, which can lead to a lower QoL. Music has been explored as a therapeutic tool to enhance QoL by promoting emotional regulation, reducing anxiety, encouraging social connections, and offering a means of self-expression. Studies show that MI, such as MT and active music engagement, can enhance mood, boost social participation, and increase overall life satisfaction in autistic individuals [22; 112].

Thompson et al. [113] conducted a randomized controlled trial on family-centered music therapy (FCMT) for young children with severe ASD, revealing notable improvements in social interactions both at home and in the community, as well as a strengthened parent-child bond. Similarly, Porter et al. [97] explored the impact of music on autistic children and adolescents, finding significant increases in self-esteem and decreases in depression scores, especially among participants aged 13 and older. Consequently, MI, particularly those that involve active participation and family engagement, can enhance social interaction and emotional well-being in ASD individuals, thereby positively impacting their QoL.

Out of 120 articles reviewed, only two [15; 22] met the criteria for our meta-analysis on the QoL in autistic individuals (**Figure 6**). The pooled effect size showed a positive trend favoring MI, with a mean difference (MD) of 4.19 [95% CI: −0.73 to 9.10], approaching statistical significance (z = 1.67, *P*-value = 0.09). This trend, while not definitive, suggests a meaningful potential for improvement in QoL through music-based therapies. Substantial heterogeneity was observed (I^2^ = 72%, *τ*^2^ = 9.16, *P*-value = 0.06), indicating considerable variability across studies, likely due to differences in outcome measures and participant characteristics, but also underscoring the diverse applicability of MI across contexts. The study by Sharda et al. [22], which used the Family Quality of Life scale (FQoL), demonstrated a significant and robust improvement (MD = 7.06, 95% CI: 2.51 to 11.61), suggesting that MT may meaningfully enhance perceived QoL in certain contexts. The study by Bieleninik et al. [15] showed a modest mean effect (MD = 2.00), but with a wider confidence interval crossing zero (95% CI: −0.70 to 4.70), reflecting more variable results. Taken together, although the meta-analysis did not reach conventional statistical significance, the overall trend is encouraging. Particularly, the strong positive results from Sharda et al. [22] support the view that MI can lead to meaningful improvements in QoL for autistic individuals, especially within family-oriented frameworks. These findings warrant further high-quality, targeted research and provide a promising foundation for incorporating MT into broader support strategies for the autistic community to improve QoL.

**Figure 6.**
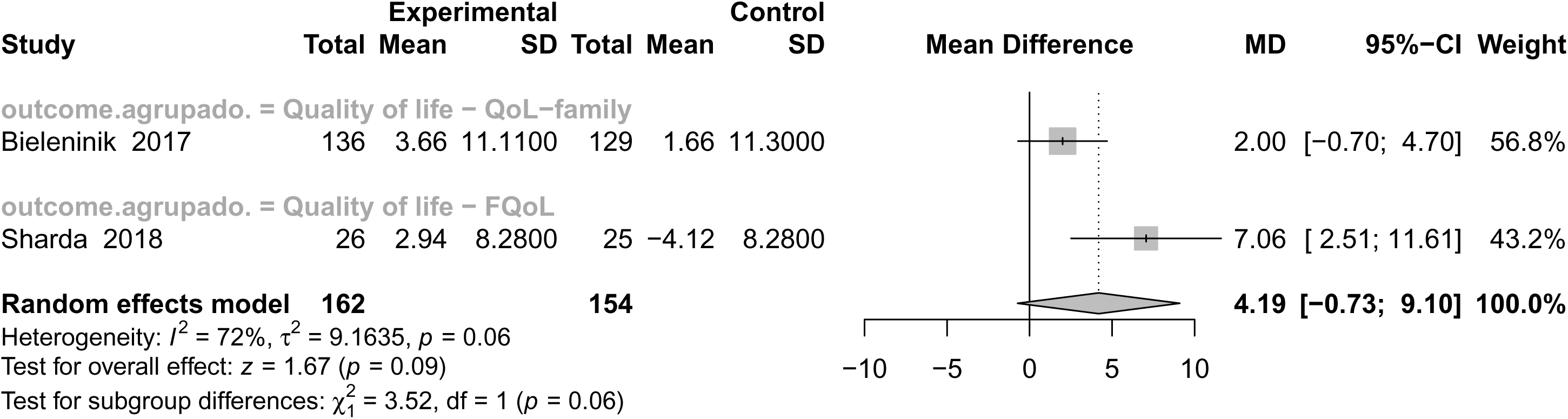
Forest plot presenting the effects of interventions on quality of life (QoL) in individuals with ASD. The figure shows mean differences between experimental and control groups, confidence intervals, and study weights. Summary estimates were generated using a random effects model, and the figure includes heterogeneity measures, prediction intervals, and subgroup/overall effect tests.

#### 3.2.5. Social interaction

Autistic individuals often face difficulties in social interaction, which can hinder their ability to build relationships and participate in shared activities. With the increasing interest in non-pharmacological interventions, MI have emerged as promising options for improving social skills in ASD individuals. The structured and predictable nature of music, along with its capacity to evoke emotional responses, makes it an effective medium for promoting social engagement. Subsequently, numerous studies have investigated the role of MI in improving social interaction in autistic individuals, providing evidence for their effectiveness in strengthening communication, emotional reciprocity, and social adaptation.

Bieleninik et al. [15] has explored the impact of MI on social interactions in autistic individuals by using ADOS-SA and reported positive outcomes. Their large-scale randomized controlled trial on MT revealed significant improvements in social communication skills among ASD children. Similarly, Ghasemtabar et al. [114] found that MT enhanced social responsiveness and emotional engagement, supporting its role in fostering meaningful social interactions. Aligning with these findings, another study [20] indicated that MT sessions improve social interaction skills in children with autism. This improvement is believed to stem from music’s ability to engage the brain’s reward systems and help regulate emotional responses, thus encouraging more positive social behaviors.

Supporting these findings, Porter et al. [97] emphasized that interactive music sessions promote social bonding and turn-taking, essential components of social interaction. Additionally, Gattino et al. [111] proved that MI led to notable improvements in social adaptation skills. Collectively, these studies suggest that MI can serve as a valuable therapeutic tool for enhancing social interaction in individuals with autism.

Out of the reviewed studies, multiple met the inclusion criteria for our meta-analysis on social interaction outcomes in individuals with ASD. Seven studies, most of which utilized the following scales: Autism Diagnosis Observation Schedule-Social Affect (ADOS-SA), Social Responsiveness Scale (SRS), and Autism Social Skills Profile (ASSP), were included in our meta-analysis assessing the effects of MI on social interaction in individuals with ASD (**Figure 7**). The pooled mean difference (MD) was 0.20, with a 95% confidence interval ranging from −0.01 to 0.42, approaching statistical significance (z = 1.89, *P*-value = 0.06). Although this result did not reach the conventional threshold for statistical significance, it indicates a small but consistent positive effect of MI on social interaction skills. The presence of moderate heterogeneity (I^2^ = 61%, *τ*^2^ = 0.04, *P*-value < 0.01) reflects differences in study populations, interventions, and, notably, outcome measurements. Most importantly, a significant test for subgroup differences (*χ*^2^ = 43.32, df = 12, *P*-value < 0.01) revealed that the observed effects varied meaningfully across the different types of social interaction outcomes assessed.

**Figure 7.**
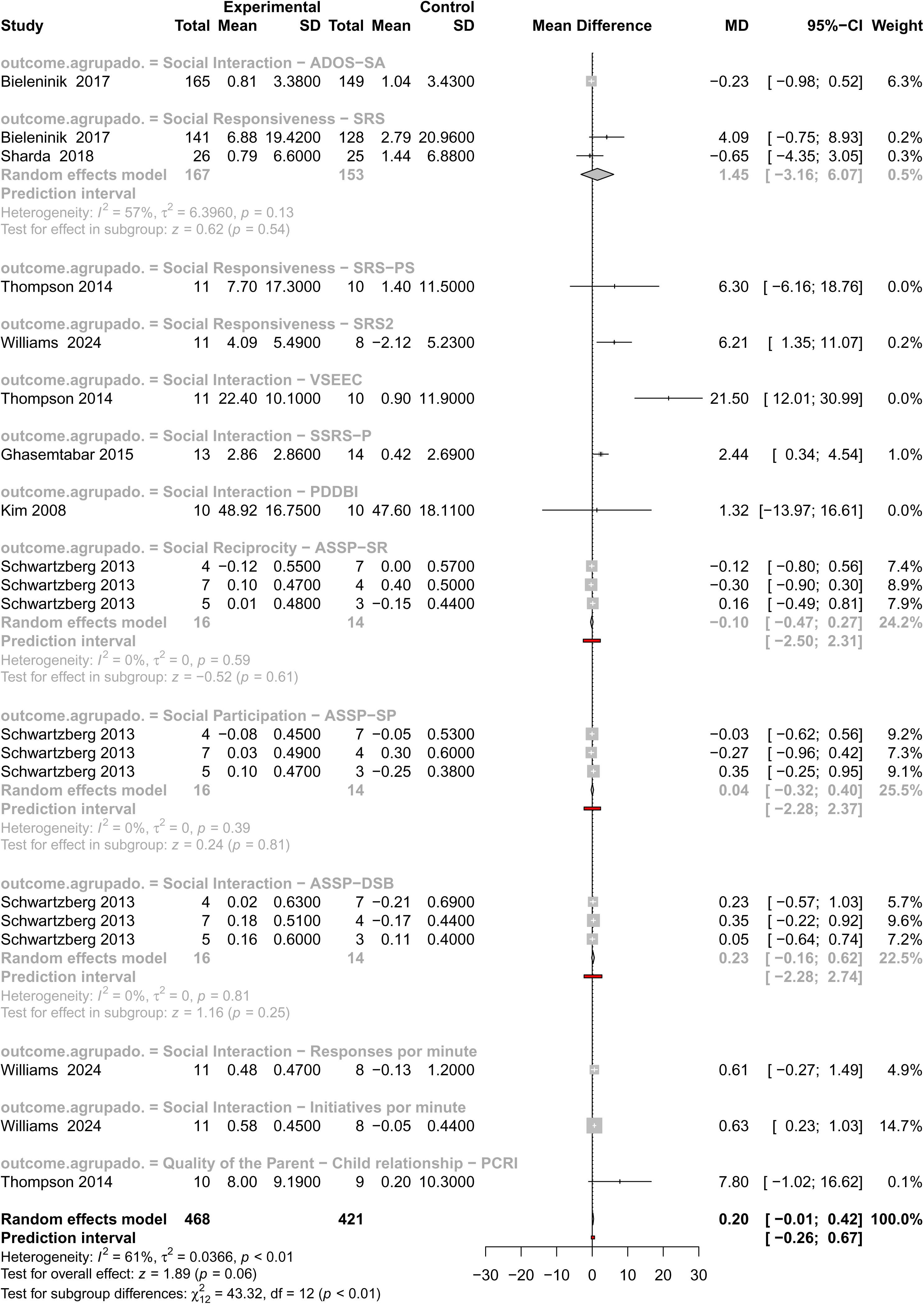
Forest plot presenting the effects of interventions on social interaction in individuals with ASD. The figure shows mean differences between experimental and control groups, confidence intervals, and study weights. Summary estimates were generated using a random effects model, and the figure includes heterogeneity measures, prediction intervals, and subgroup/overall effect tests.

The studies encompassed a wide range of validated tools, each targeting distinct dimensions of social functioning. The ADOS-SA subgroup used the Autism Diagnostic Observation Schedule, Social Affect domain, which captures clinician-observed core deficits in social communication. In contrast, the Social Responsiveness – SRS, SRS-PS (preschool version), and SRS-2 subgroups relied on the Social Responsiveness Scale in its various forms, a widely adopted parent-or teacher-report instrument designed to measure the severity of autism-related social difficulties in everyday life. Other subgroups focused on different developmental and relational aspects of social interaction. For example, the VSEEC group used the Vineland Social-Emotional Early Childhood Scales to assess emotional and social functioning in young children, while the SSRS-P subgroup employed the Social Skills Rating System – Parent form to evaluate caregiver perceptions of social behavior. The Pervasive Developmental Disorders Behavior Inventory (PDDBI) offers a broader behavioral assessment that includes communication and adaptive functioning. Several subgroups drew on subdomains of the Assessment of Social and Sensory Participation (ASSP). These included Social Reciprocity – ASSP-SR, which reflects mutual, responsive social behaviors; Social Participation – ASSP-SP, capturing engagement in structured and unstructured social contexts; and ASSP-DSB, which focuses on daily social behavior. In addition to these standardized assessments, some studies measured more granular behavioral outcomes such as “Responses per minute” and “Initiatives per minute”, derived from observational coding of spontaneous social behaviors in naturalistic settings. Lastly, the Parent–Child Relationship – PCRI subgroup used the Parent–Child Relationship Inventory to assess perceived relational quality, including communication and emotional closeness between caregiver and child.

Among the various subgroups analyzed, only three demonstrated statistically significant improvements in social interaction outcomes following MI. The VSEEC subgroup showed the most substantial effect, with a mean difference (MD) of 21.50 and a 95% CI of [12.01; 30.99], indicating a large and robust improvement in social-emotional functioning in young children. Similarly, the SRS2 subgroup showed a significant positive effect (MD = 6.21, 95% CI: [1.35; 11.07]). Additionally, the SSRS-P subgroup revealed a moderate but meaningful improvement (MD = 2.44, 95% CI: [0.34; 4.54]), suggesting increased caregiver-observed social engagement.

These findings suggest that MI may be particularly effective when assessed using tools that capture real-world, functional, and caregiver-sensitive aspects of social behavior, rather than more rigid or clinical instruments. The consistency of these effects across observational and parent-report measures underscores the practical relevance and ecological validity of music-based therapies in supporting social development in ASD.

Taken together, these results are promising. Although the overall meta-analytic effect narrowly missed statistical significance, the consistent positive direction of the effects, combined with substantial and meaningful subgroup differences, indicates that MI can foster improvements in social interaction among autistic individuals, particularly when targeted to specific contexts and measured through responsive, ecologically valid tools. These findings emphasize the importance of selecting appropriate outcome measures and tailoring interventions to individual needs, and they justify further high-quality, stratified research in this area.

## 4. Discussion

Research on musical processing and abilities in individuals with ASD reveals a distinctive cognitive profile, often described as a potential “musical phenotype”. Among 45 studies, findings consistently show that many autistic individuals, even without formal musical training, demonstrate enhanced sensitivity to pitch changes, superior pitch discrimination, and in some cases, a higher prevalence of absolute pitch compared to the general population. These abilities appear to stem from atypical cognitive styles, including heightened perceptual functioning, focused attention, and in some cases, superior memory for musical material. Although musical processing in ASD is often preserved, certain aspects, such as pitch production or emotional expressivity, can vary, with some impairments emerging particularly in speech contexts. Emotional responses to music in ASD are generally comparable to those of TD individuals, though differences in physiological arousal, processing speed, and neural activation patterns suggest unique underlying mechanisms. Neuroimaging studies further support these distinctions, indicating that while basic emotional recognition remains intact, brain regions involved in timing, coordination, and autonomic regulation may function differently during musical experiences in ASD [77]. Studies also show that individuals with ASD experience strong preferences for certain types of music, with preferences shaped by both sensory sensitivity and therapeutic utility. For instance, simpler musical structures (e.g., predictable rhythms and consonant harmonies) appear particularly effective in promoting attention and reducing stereotypical behaviors, especially in individuals with more severe ASD symptoms [22; 76]. Qualitative research emphasizes the importance of music in emotional self-regulation, identity formation, and social connection, reinforcing its role as a meaningful and supportive element in the lives of people with ASD. Together, these results highlight music’s potential as a powerful tool for engagement, communication, and personal expression within this population.

Our investigation aimed to evaluate the effectiveness of MI in ASD populations. The synthesis encompassed a wide range of outcomes, including attention, verbal and non-verbal communication, QoL, behavior, and social interaction, assessed using various standardized measures. The findings reveal a complex pattern of results, with some domains exhibiting promising effects and others showing limited or non-significant improvements. Notably, the heterogeneity observed both across and within outcome domains reflects variability in study designs, intervention types, measurement tools, and participant characteristics. Overall, the results suggest that MI has a moderate, positive effect on several developmental outcomes in individuals with ASD, supporting its value as a therapeutic approach. The results of our meta-analysis provide a noteworthy contribution when compared to prior systematic reviews in the field of ASD and MI. This is primarily due to the more rigorous methodological framework we applied, which included clearly defined inclusion and exclusion criteria that carefully considered participant characteristics, intervention types, and study design. Our review also encompassed a broader range of participant ages, timeframes, and outcome domains, enhancing the comprehensiveness and applicability of the findings. Crucially, we combined both qualitative and quantitative analytical approaches, which allowed for a more balanced and context-sensitive interpretation of results, particularly given the diversity of outcome measures used across studies. By focusing on different outcomes that hold direct relevance for individuals with ASD, this work adds meaningful depth to the existing body of literature.

A total of 15 trials met the criteria for inclusion in the meta-analysis, evaluating the effects of MI on ASD individuals ranging from two years of age to young adulthood. These interventions were compared against standard care or placebo-like control therapies designed to isolate the unique contribution of music itself by controlling for non-specific factors such as therapist attention and participant engagement. Our meta-analyses aimed to evaluate the effectiveness of MI, including music-making, receptive music listening, and MT, across several developmental domains in individuals with ASD, including attention, behavior, verbal and non-verbal communication, QoL, and social interaction. Overall, the findings present a complex picture: while statistically significant effects were not consistently observed across all domains, multiple positive trends emerged, particularly within specific subgroups and outcome measures [90; 92; 96; 115]. These results highlight both the therapeutic potential of MI, and the complexity involved in evaluating its efficacy in autistic populations. The findings align with theoretical perspectives suggesting that music may engage neural systems associated with emotion, language, and social cognition, domains that are typically affected in individuals with ASD [23].

Across the attention domain, the pooled results did not yield statistically significant effects. However, a small positive trend was observed, particularly after removing a single outlier from the analysis, which strengthened the results and suggests that music may modestly influence attentional processes. High heterogeneity across the included studies highlights the variation in measurement tools, intervention formats, and participant characteristics, suggesting that the potential for MI to enhance attention may depend heavily on the type of attentional skill targeted and the contextual features of the intervention. Given that musical experiences often require sustained focus and dynamic engagement, further exploration is warranted, particularly with refined methodologies and attention subtypes more closely aligned with musical structures. Behavioral outcomes showed no significant overall effect, although individual studies yielded mixed findings. Some reported meaningful improvements, while others observed null or inconsistent effects. Notably, removing a single outlier from the meta-analysis resulted in a more consistently positive outcome, reaching marginal statistical significance. The extremely high heterogeneity across studies, driven by differences in behavioral scales, intervention types, and participant characteristics, suggests that the broad category of’behavior’ may be too general to capture the specific domains in which MI might be most effective. Rather than expecting uniform behavioral changes, future research should focus on targeted areas such as emotional regulation, repetitive behaviors, or adaptive functioning, using standardized and validated assessment tools. In terms of verbal communication, the pooled analysis across various outcome measures again failed to reveal a statistically significant effect. However, a closer examination of subgroups uncovered more promising findings. Notably, MI appeared to enhance expressive language abilities, particularly at the phonemic level, such as vowel articulation, where the effects reached statistical significance. These findings are consistent with the natural overlap between musical and linguistic processing, especially regarding rhythm, pitch, and prosody. The capacity of music to structure and scaffold auditory input may thus support specific speech production skills in individuals with ASD. This domain exemplifies the importance of fine-grained outcome analysis, as global language measures may obscure more focal gains. The analysis of non-verbal communication outcomes did not yield significant effects either, though small positive trends were observed. Interestingly, heterogeneity in this domain was relatively low, which may suggest a more consistent, albeit modest, impact of MI. Individual studies employing sensitive observational tools reported slight improvements, indicating that MI might influence non-verbal communicative behaviors such as gestures, eye contact, or joint attention, particularly when interventions are developmentally attuned and interaction focused. In contrast, the results related to QoL were more encouraging, despite being drawn from only two studies. The pooled effect approached statistical significance, and one study demonstrated a robust improvement in family-related QoL outcomes. This points to the broader psychosocial benefits of MT, which may extend beyond individual-level symptom change to influence familial and relational well-being. These findings support a shift toward evaluating MI not solely on clinical symptoms, but on broader, person-centered outcomes that reflect real-world functioning and QoL. Among all domains, social interaction showed the most consistently positive pattern. Although the overall effect narrowly missed statistical significance, multiple subgroups, especially those relying on caregiver reports or ecologically valid observational tools, demonstrated significant improvements. Instruments such as the VSEEC, the SRS-2, and the SSRS-P captured meaningful changes in social behaviors, suggesting that MI may enhance social engagement, reciprocity, and functional interaction in everyday contexts.

Our findings seem to support the notion that MI outperforms standard care and has therapeutic properties. The musical experience itself, especially when grounded in relational dynamics and shaped by the individual’s interests and motivations, appears to foster social engagement in ways that more traditional therapies may not. These approaches have shown promise in enhancing core social communication skills, such as initiating interactions, maintaining eye contact, and interpreting emotional stimuli [111].

Despite the promising outcomes identified in this review, several methodological and conceptual limitations should be acknowledged. A major restriction lies in the relatively small sample sizes of many included studies, which diminish statistical power and limit the generalizability of the findings across the heterogeneous autism spectrum. Future research would greatly benefit from larger, more representative cohorts that account for variability in age, language ability, and cognitive profiles. Additionally, most studies investigated only short-term effects, making it difficult to assess whether observed gains in communication, attention, behavior, or QoL are sustained over time. The lack of longitudinal follow-up data hinders our understanding of the long-term efficacy and developmental trajectory of MI, which is particularly relevant in neurodevelopmental conditions such as autism. Another key limitation concerns the ecological validity of the outcome measures used to evaluate the impact of MI. As highlighted by Heaton et al. [43], musical assessments often rely on controlled tasks that do not adequately capture the complexity of real-world musical experiences. Some experimental paradigms focus narrowly on isolated musical features, such as pitch or rhythm discrimination, without considering how music is experienced dynamically, socially, and emotionally in everyday life. This methodological gap may lead to underestimations of music’s broader impact on engagement, emotional regulation, and social connection. Moreover, standardized tests may fail to reflect individual preferences, sensitivity, and the highly personalized nature of musical engagement in autistic individuals [81].

Therefore, to advance the field, future research should prioritize longitudinal designs that can track outcomes over extended periods and across real-world settings. There is also a need for greater attention to the ecological validity of outcome measures, incorporating assessments that capture spontaneous musical interaction and meaningful engagement. Understanding the specific musical abilities and processing profiles of individuals with ASD is essential for developing neurobiologically grounded, personalized interventions. Moreover, the incorporation of biological data, such as genetic, epigenetic, or transcriptomic information, into intervention studies (in line with other disease conditions, see [116; 117; 118]) could provide a deeper understanding of the underlying mechanisms and help identify biomarkers predictive of treatment response. Integrating behavioral outcomes with molecular and neurophysiological measures represents a promising direction for future research [101].

Despite the well-documented difficulties that individuals with ASD may encounter when processing complex emotional or social indicators, numerous studies have demonstrated that the recognition of basic emotions in music is often preserved. This suggests that music constitutes a domain of both relative cognitive strength and powerful intrinsic interest. As a non-verbal, emotionally resonant medium, music offers an alternative channel of communication that is particularly valuable in populations with language impairments.

Taken together, the findings of this meta-analysis suggest that while MI may not yield uniformly large or statistically robust effects across all developmental domains in ASD, they show meaningful promise when applied in a targeted, individualized, and context-sensitive manner. The considerable heterogeneity across studies, stemming from differences in intervention formats (e.g., active *vs*. passive engagement, session length, therapist involvement), participant characteristics (e.g., age, language ability, ASD severity), and outcome measures complicates broad generalizations but also highlights the flexibility of MI to meet diverse needs. Notably, emerging patterns of benefit in areas such as expressive language, phonemic articulation, and social interaction point to the promising capacity of music to promote engagement and foster meaningful connection. As an empirically grounded and neurodiversity-affirming tool, MI represents a compelling complement to conventional therapeutic approaches. Future research should explore long-term effects, skill generalization, and the role of individual differences in treatment response to better integrate MI into personalized, responsive, and holistic care for autistic individuals.

## Data Availability

This is a systematic review and a meta-analysis. It uses data publicly available.

## Acknowledgements

The authors would like to express their appreciation to the study investigators of the Sensogenomics network (sensogenomics.com; Sensogenomics Working Group [see Annex]), as well as the nursery and laboratory service at the Hospital Clínico Universitario de Santiago de Compostela, for their invaluable dedication and support. This work was supported by: *i*) GAIN IN607B 2020/08 and IN607A 2023/02, and EUTERPE_adn (Programa de Cooperación Interreg-VI POCTEP; Ref. 0313_EUTERPE_ADN_1_E) (to A.S.), and IIN607A2021/05 (to F.M.-T.), and *ii*) Consorcio Centro de Investigación Biomédica en Red de Enfermedades Respiratorias (CB21/06/00103; to A.S. and F.M.-T.). The funders were not involved in the study design, collection, analysis, interpretation of data, the writing of this article, or the decision to submit it for publication.

## Supplementary Data

Figure S1. Forest plot illustrating the effect sizes (mean differences) for studies comparing intervention and control groups on attention outcomes in individuals with ASD, excluding the study by Sá et al. [90] from the meta-analysis.

Figure S2. Forest plot illustrating the effect sizes (mean differences) for studies comparing intervention and control groups on attention outcomes in individuals with ASD, excluding the RBS-R Sensory subscale results from Srinivasan et al. [96] from the meta-analysis.

## Sensogenomics Working Group

Antonio Salas Ellacuriaga – PI; Federico Martinón-Torres – PI; Laura Navarro Ramón – Coordinator

### GenPoB/GenVip - Instituto de Investigación Sanitaria (IDIS) (alphabetic order)

Alba Camino Mera, Albert Padín Villar, Alberto Gómez Carballa, Alejandro Pérez López, Alicia Carballal Fernández, Ana Cotovad Bellas, Ana Isabel Dacosta Urbieta, Narmeen Mallah, Ana María Pastoriza Mourelle, Ana María Senín Ferreiro, Andrés Muy Pérez, Antía Rivas Oural, Antonio Justicia Grande, Antonio Piñeiro García, Anxela Cristina Delgado García, Belén Mosquera Pérez, Blanca Díaz Esteban, Carlos Durán Suárez, Carmen Curros Novo, Carmen Gómez Vieites, Carmen Rodríguez-Tenreiro Sánchez, Celia Varela Pájaro, Claudia Navarro Gonzalo, Cristina Serén Trasorras, Cristina Talavero González, Einés Monteagudo Vilavedra, Estefanía Rey Campos, Esther Montero Campos, Fernando Álvez González, Fernando Caamaño Viñas, Francisco García Iglesias, Gloria Viz Rodríguez, Hugo Alberto Tovar Velasco, Irene Álvarez Rodríguez, Irene García Zuazola, Irene Rivero Calle, Iria Afonso Carrasco, Isabel Ferreirós Vidal, Isabel Lista García, Isabel Rego Lijo, Iván Prieto Gómez, Iván Quintana Cepedal, Jacobo Pardo Seco, Jesús Eirís Puñal, José Gómez Rial, José Manuel Fernández García, José María Martinón Martínez, Julia Cela Mosquera, Julia García Currás, Julián Montoto Louzao, Lara Martínez Martínez, Laura Navarro Marrón, Lidia Piñeiro Rodríguez, Lorenzo Redondo Collazo, Lúa Castelo Martínez, Lucía Company Arciniegas, Luis Crego Rodríguez, Luisa García Vicente, Manuel Vázquez Donsión, María Dolores Martínez García, María Elena Gamborino Caramés, María Elena Sobrino Fernández, María José Currás Tuala, María Martínez Leis, María Soledad Vilas Iglesias, María Sol Rodriguez Calvo, María Teresa Autran García, Marina Casas Pérez, Marta Aldonza Torres, Marta Bouzón Alejandro, Marta Lendoiro Fuentes, Miriam Ben García, Miriam Cebey López, Montserrat López Franco, Nour El Zahraa Mallah, Narmeen Mallah, Natalia García Sánchez, Natalia Vieito Perez, Patricia Regueiro Casuso, Ricardo Suárez Camacho, Rita García Fernández, Rita Varela Estévez, Rosaura Picáns Leis, Ruth Barral Arca, Sandra Carnota Antonio, Sandra Viz Lasheras, Sara Pischedda, Sara Rey Vázquez, Sonia Marcos Alonso, Sonia Serén Fernández, Susana Rey García, Vanesa Álvarez Iglesias, Victoria Redondo Cervantes, Vanesa Álvarez Iglesias, Wiktor Dominik Nowak, Xabier Bello Paderne, Xabier Mazaira López

### Nursing volunteers (alphabetic order)

Alejandra Fernández Méndez, Ana Isabel Abadín Campaña, Ana María León Caamaño, Ana María Buide Illobre, Ángeles Mera Cores, Carmen Nieves Vastro, Carolina Suarez Crego, Concepción Rey Iglesias, Cristina Candal Regueira, Dolores Barreiro Puente, Elvira Rodríguez Rodríguez, Eugenia González Budiño, Eva Rey Álvarez, Fernando Rodríguez Gerpe, Gemma Albela Silva, Isabel Castro Pérez, Isabel Domínguez Ríos, José Ángel Fernández de la Iglesia, José Cruces Vázquez, José Luis Cambeiro Quintela, José Ramón Magariños Iglesias, Julia Rey Brandariz, Julio Abel Fernández López, Luisa García Vicente, Manuel González Lito, Manuel González Lijó, Manuela Pérez Rivas, Margarita Turnes Paredes, María Aurora Méndez López, María Begoña Tomé Arufe, María Campos Torres, María del Carmen Baloira Nogueira, María del Carmen García juan, María Esther Moricosa García, María Luz Chao Jarel, María Martínez Leis, María Mercedes Jiménez Santos, María Salomé Buide Illobre, María Victoria López Pereira, Mercedes Jorge González, Mercedes Isolina Rodríguez Rodríguez, Miren Payo Puente, Natalia Carter Domínguez, Olga María Reyes González, Pilar Mera Rodríguez, Purificación Sebio Brandariz, Salomé Quintáns lago, Yolanda Rodríguez Taboada, María Pereira Grau.

### Other volunteers (alphabetic order)

Alba Arias Gómez, Alejandro Moreno Díaz, Ana Arca Marán, Astro González Guirado, Brais García Iglesias, Carlos Sánchez Rubín, Carmen Otero de Andrés, Clara Pérez Errazquin Barrera, Claudia Rey Posse, Cristina Rojas García, Eduardo Xavier Giménez Bargiela, Elena Gloria Morales García, Fabio Izquierdo García Escribano, Gabriel Guisande García, Jaime López Martín, Lara Pais Ramiro, Lucía Rico Montero, Luís Estévez Martínez, Manuel Estévez Casal, María Aránzazu Palomino Caño, María Rubio Valdés, Marisol Nogales Benítez, Miryam Tilve Pérez, Nuria Villar Muiños, Pablo Del Cerro Rodríguez, Pablo Pozuelo Martínez Cardeñoso, Salma Ouahabi El Ouahabi, Santiago Vázquez Calvache

