## Supplementary material for "The effect of music interventions in autism spectrum disorder: A systematic review and meta-analysis": Figure S1

| Study | Experimental Total | Experimental Mean | Experimental SD | Control Total | Control Mean | Control SD | MD | 95%−CI | Weight |
|---|---|---|---|---|---|---|---|---|---|
| **Outcome = Attention − TEA−Ch2** | | | | | | | | | |
| Sa 2020 | 11 | −4.40 | 11.7900 | 12 | −6.00 | 11.2500 | 1.60 | [ −7.84; 11.04] | 19.7% |
| **Outcome = Attention − RBBS** | | | | | | | | | |
| Sa 2020 | 11 | 3.37 | 2.7500 | 12 | 2.66 | 1.6600 | 0.71 | [ −1.17; 2.59] | 35.7% |
| **Outcome = Joint Attention − With child** | | | | | | | | | |
| LaGasse 2014 | 9 | 8.23 | 11.4500 | 8 | −6.63 | 7.4400 | 14.86 | [ 5.77; 23.95] | 20.4% |
| **Outcome = Joint Attention − With adults** | | | | | | | | | |
| LaGasse 2014 | 9 | −1.61 | 6.3600 | 8 | −1.81 | 8.8100 | 0.20 | [ −7.18; 7.58] | 24.1% |
| **Random effects model** | **40** | | | **40** | | | **3.66** | **[ −2.49; 9.80]** | **100.0%** |
| **Prediction interval** | | | | | | | | **[−22.31; 29.62]** | |

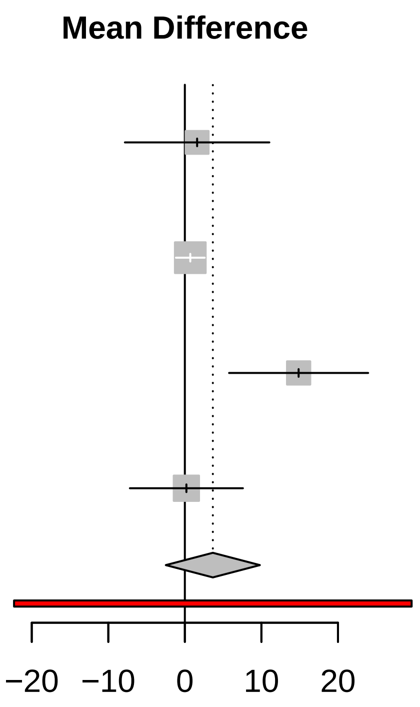


Heterogeneity: $I^2 = 67\%$, $\tau^2 = 26.5984$, $p = 0.03$
Test for overall effect: $z = 1.17$ ($p = 0.24$)
Test for subgroup differences: $\chi^2_3 = 9.02$, df = 3 ($p = 0.03$)
