## Supplementary material for "The effect of music interventions in autism spectrum disorder: A systematic review and meta-analysis": Figure S2

| Study | Experimental Total | Experimental Mean | Experimental SD | Control Total | Control Mean | Control SD | MD | 95%−CI | Weight |
|---|---|---|---|---|---|---|---|---|---|
| **Outcome = Behavior − VABS−MB** | | | | | | | | | |
| Sharda 2018 | 26 | 0.38 | 0.4300 | 24 | 0.46 | 0.5200 | −0.08 | [ −0.35; 0.19] | 16.7% |
| **Outcome = Behavior − VABS−3** | | | | | | | | | |
| Williams 2024 | 11 | 2.27 | 3.4900 | 8 | 1.12 | 2.9800 | 1.15 | [ −1.77; 4.07] | 16.3% |
| **Outcome = Behavior − RBS−R−Negative** | | | | | | | | | |
| Srinivasan 2015 | 12 | 33.10 | 30.7900 | 12 | −6.80 | 11.7100 | 39.90 | [ 21.26; 58.54] | 8.3% |
| **Outcome = Behavior − RBS−R−Stereotypes** | | | | | | | | | |
| Srinivasan 2015 | 12 | −1.40 | 12.8700 | 12 | −2.10 | 8.1500 | 0.70 | [ −7.92; 9.32] | 13.8% |
| **Outcome = Behavior − CARS** | | | | | | | | | |
| Ren 2022 | 15 | 5.18 | 0.6900 | 15 | 0.70 | 0.7400 | 4.48 | [ 3.97; 4.99] | 16.7% |
| **Outcome = Behaviour − Eye gaze** | | | | | | | | | |
| LaGasse 2014 | 9 | 3.73 | 15.6600 | 8 | −14.75 | 10.0300 | 18.48 | [ 6.11; 30.85] | 11.6% |
| **Outcome = Behaviour − Global improvement−ATEC** | | | | | | | | | |
| Ren 2022 | 15 | 5.76 | 2.4100 | 15 | 0.03 | 3.0900 | 5.73 | [ 3.75; 7.71] | 16.5% |
| **Random effects model** | **100** | | | **94** | | | **7.43** | **[ −0.16; 15.03]** | **100.0%** |
| **Prediction interval** | | | | | | | | **[−18.88; 33.74]** | |

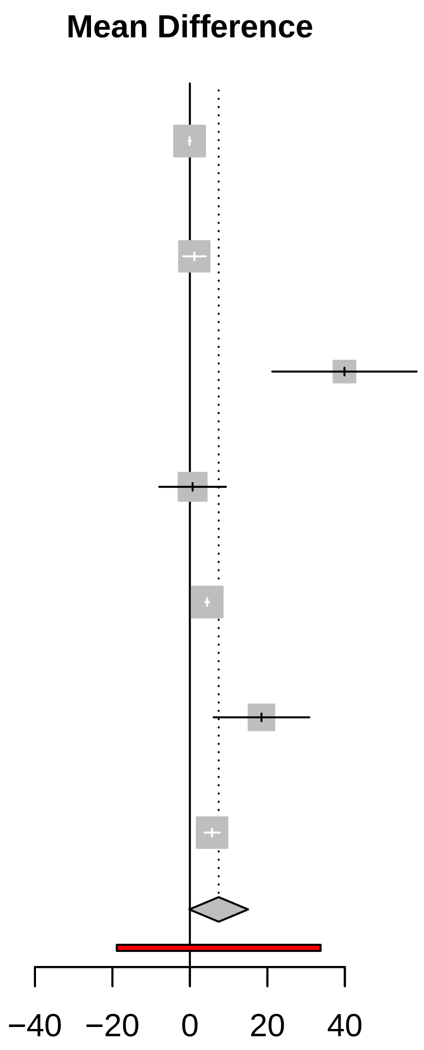


Heterogeneity: $I^2 = 98\%$, $\tau^2 = 89.7428$, $p < 0.01$
Test for overall effect: $z = 1.92$ ($p = 0.06$)
Test for subgroup differences: $\chi^2_6 = 287.12$, df = 6 ($p < 0.01$)
